# *MTHFR* C677T polymorphism and promoter methylation in schizophrenia patients with type 2 diabetes mellitus: evidence from a Han Chinese cohort

**DOI:** 10.64898/2026.04.09.26350471

**Authors:** Cuncun Yang, Rongrong Li, Xizi Wang, Kun Li, Fengjiao Yuan, Xiaodong Jia, Rui Zhang, Juan Zheng

## Abstract

Schizophrenia (SCZ) and type 2 diabetes mellitus (T2DM) are common comorbid disorders that severely impair patient prognosis and quality of life. This study aimed to explore the association between the methylenetetrahydrofolate reductase (*MTHFR*) C677T gene polymorphism and *MTHFR* promoter methylation in patients with comorbid SCZ and T2DM. A total of 120 participants were enrolled from Liaocheng Fourth People’s Hospital between January 2025 and June 2025, comprising 30 subjects in each of the four groups: SCZ group, T2DM group, SCZ–T2DM comorbid (SCZ+T2DM) group, and healthy control (CTL) group. Corresponding primers were designed for genetic analysis, and methylation-specific PCR (MSP) was performed to detect the methylation level of the *MTHFR* promoter. Genotype distribution of the *MTHFR* C677T polymorphism was consistent with Hardy–Weinberg equilibrium (HWE) (*p*>0.05). The C677T polymorphism was significantly associated with an elevated risk of SCZ and T2DM comorbidity (*p*<0.05). Notably, the methylation rate of the *MTHFR* promoter in the SCZ+T2DM group (95.00%) was not significantly higher than that in the CTL group (90.00%) (*p*>0.05). In conclusion, the *MTHFR* gene may serve as a susceptibility gene for SCZ–T2DM comorbidity, whereas *MTHFR* promoter methylation is not associated with the pathogenesis of this comorbid condition. These results indicate that genetic variation in *MTHFR*, rather than promoter methylation, contributes critically to the comorbidity of SCZ and T2DM in the Han Chinese population. Our findings may provide novel molecular insights into their shared pathophysiology and inform future clinical strategies for patients with this complex phenotype.

## 1 Introduction

Schizophrenia (SCZ) is a multifaceted disorder characterized by chronic psychosis, cognitive impairment, and functional disability[1], representing a major global public health concern[2]. It can be categorized into several subtypes, including simple, catatonic, paranoid, hebephrenic, and undifferentiated forms[3]. Current epidemiological data show that more than 23 million people worldwide are affected by SCZ, corresponding to 1% of the global population, with a prevalence ranging from 0.5% to 0.8%[4]. In China, nearly 10 million individuals have been diagnosed with SCZ, with a prevalence rate of 0.7%, and this figure continues to increase annually[5]. Studies conducted in the United States reveal a 1.3% lifetime prevalence of schizophrenia, a disorder that follows a persistent lifelong course once diagnosed[6]. The economic cost of SCZ management in the United States accounts for 0.8% of the gross domestic product, underscoring its substantial financial impact on healthcare systems[7]. Beyond clinical manifestations, SCZ imposes profound suffering on patients and their families, while also placing a heavy socioeconomic burden on society[8]. Despite extensive research efforts, the exact etiology of SCZ remains incompletely unclear; however, it is generally accepted as a complex disorder driven by a combination of genetic and environmental factors.

Type 2 diabetes mellitus (T2DM) is a highly prevalent and severe chronic metabolic disorder characterized by inadequate insulin secretion, which results in chronic hyperglycemia[9]. This pathological state can give rise to a spectrum of metabolic complications, including cardiovascular diseases, diabetic retinopathy (DR), diabetic nephropathy (DN), and diabetic peripheral neuropathy (DPN)[10]. T2DM is primarily manifested as systemic disturbances in glucose, lipid, and protein metabolism, which may further contribute to the dysfunction of multiple organs. T2DM not only compromises physical and mental health but also impairs the quality of life and shortens the life expectancy of affected individuals[11]. According to data from the International Diabetes Federation (IDF), the global prevalence of diabetes was approximately 9.3% (463 million individuals) in 2019, and is projected to rise to 10.2% (578 million) by 2030 and 10.9% (700 million) by 2045[12]. In China, approximately 116 million individuals are living with diabetes, of whom more than 95% are diagnosed with T2DM[13]. Accumulating evidence has demonstrated that individuals with schizophrenia are 2–5 times more likely to develop T2DM compared with the general population, highlighting the substantial public health challenges posed by the comorbidity of SCZ and T2DM[14]. Although the etiological mechanisms underlying both schizophrenia and T2DM remain incompletely elucidated, both disorders are recognized as genetically heterogeneous polygenic diseases modulated by the interplay of genetic and environmental factors.

Methylenetetrahydrofolate reductase (MTHFR) is a pivotal enzyme in folate metabolism, responsible for catalyzing the irreversible conversion of 5,10-methylenetetrahydrofolate to 5-methyltetrahydrofolate[15]. This product acts as the major methyl donor for the remethylation of homocysteine to methionine[16]. The *MTHFR* gene is localized on chromosome 1p36.3 and harbors a common single-nucleotide polymorphism (SNP), C677T (rs1801133), which gives rise to an alanine-to-valine substitution at codon 222[17]. This amino acid replacement impairs MTHFR enzymatic activity and may consequently result in elevated homocysteine levels (hyperhomocysteinemia)[18]. Previous meta-analyses have revealed significant associations between the *MTHFR* C677T polymorphism and an increased susceptibility to both SCZ and T2DM in Asian populations. For instance, a meta-analysis comprising 81 studies[19]demonstrated a significant relationship between the *MTHFR* C677T polymorphism and SCZ risk in Asian populations, while another meta-analysis of 68 studies[20]identified an association between this variant and T2DM. Collectively, these findings suggest that the *MTHFR* C677T polymorphism may serve as a shared genetic risk factor for both SCZ and T2DM.

Epigenetic modifications, especially DNA methylation, play a critical role in the onset and progression of complex diseases by regulating gene expression without altering the underlying DNA sequence[21]. *MTHFR* promoter methylation can suppress gene transcription, decrease MTHFR expression and enzymatic activity, and further disrupt folate metabolism and homocysteine (Hcy) homeostasis[22–23].Previous studies[24–25]have demonstrated that hypermethylation of the *MTHFR* promoter is associated with diabetic complications, including retinopathy and nephropathy, and that the *MTHFR* C677T polymorphism may interact with DNA methylation to modulate susceptibility to neuropsychiatric disorders. Nevertheless, few studies have simultaneously investigated the contributions of both the *MTHFR* C677T polymorphism and promoter methylation to the comorbidity of SCZ and T2DM, particularly within the Han Chinese population.

Based on the aforementioned research, we propose the scientific hypothesis that schizophrenia and type 2 diabetes mellitus share common genetic and epigenetic regulatory mechanisms, which represents a key research direction for understanding their comorbidity. We also aim to clarify whether the methylation status of CpG islands within the *MTHFR* promoter region acts as a susceptibility factor affecting gene expression and enzymatic activity. Thus, this study will systematically verify this hypothesis and explore the potential effects of *MTHFR* polymorphisms and related methylation patterns in patients comorbid with SCZ and T2DM. Collectively, this work aims to provide novel insights into the genetic and epigenetic mechanisms underlying SCZ–T2DM comorbidity, and may contribute to the development of targeted prevention and intervention strategies for high-risk populations.

## 2 Materials and methods

### 2.1 Subjects

All participants were recruited as inpatients from the Liaocheng Fourth People’s Hospital, a specialized psychiatric hospital in Shandong Province, China, between January 2025 and June 2025. A total of 120 subjects were enrolled and divided into four groups (n = 30 per group): schizophrenia only (SCZ), type 2 diabetes mellitus only (T2DM), comorbid SCZ and T2DM (SCZ+T2DM), and healthy controls (CTL). A 5- mL fresh peripheral blood sample was collected from each participant. For the SCZ group, the inclusion criteria were as follows: (1) Han Chinese individuals diagnosed with schizophrenia according to the Diagnostic and Statistical Manual of Mental Disorders, 5th Edition (DSM- 5) criteria[26]; (2) illness duration of at least 2 years; (3) stable doses of oral antipsychotic medications for at least 6 months prior to enrollment. Exclusion criteria included: (1) psychotic disorders attributable to other somatic diseases (neurological or non- neurological); (2) comorbid psychiatric disorders other than schizophrenia. For the T2DM group, inclusion criteria were:(1) Han Chinese individuals diagnosed with T2DM according to the 2003 clinical diagnostic criteria of the American Diabetes Association (ADA); (2) fasting plasma glucose (FPG) ≥ 7.0 mmol/L; or (3) 2- hour postprandial plasma glucose (2hPG) ≥ 11.1 mmol/L. Patients with type 1 diabetes, gestational diabetes, or other specific types of diabetes were excluded. The SCZ+T2DM group comprised patients who met both the DSM- 5 criteria for schizophrenia and the 2003 ADA criteria for T2DM, with FPG ≥ 7.0 mmol/L and 2hPG ≥ 11.1 mmol/L. Exclusion criteria were: (1) mental disorders induced by psychoactive substances or non- addictive substances; (2) organic mental disorders; (3) a history of neurological diseases or other major medical illnesses; (4) malignant tumors; (5) other brain disorders; (6) pregnancy or lactation. Healthy controls were recruited from hospital staff and voluntary blood donors. All controls were physically healthy, with no severe chronic diseases and no family history of psychiatric disorders, as assessed by a psychiatrist using the Mini International Neuropsychiatric Interview. The study protocol was approved by the Institutional Ethics Committee of Liaocheng Fourth People’s Hospital. Written informed consent was obtained from all participants prior to enrollment, and all procedures were performed in accordance with the ethical principles outlined in the Declaration of Helsinki.

### 2.2 Genotyping

Genomic DNA was extracted from 200 μL of peripheral whole blood using the TIANamp Genomic DNA Kit (Tiangen, Beijing, China), following the manufacturer’s protocol. DNA purity was evaluated by measuring the absorbance at 260 nm and 280 nm; samples with an A260/A280 ratio ranging from 1.8 to 2.0 were deemed acceptable for subsequent experiments. Genotyping of the *MTHFR* C677T polymorphism was performed using allele-specific polymerase chain reaction (PCR) with the following primers: forward 5′-CCCAGTCCCTGTGGTCTCTT-3′, reverse 5′-ACTCAGCACTCCACCCAGAG-3′. Each 25 μL PCR reaction consisted of 2.5 μL DNA template, 1.25 μL of each primer, 12.5 μL of 2× San Taq PCR Master Mix (Sangon Biotech, Shanghai, China), and 7.5 μL nuclease-free deionized distilled water (ddH₂O). PCR amplification was performed under the following conditions: initial denaturation at 94°C for 5 min; 35 cycles of denaturation at 94°C for 30 s, annealing at 62°C for 30 s, and extension at 72°C for 30 s; followed by a final extension at 72°C for 10 min. The amplified 366 bp PCR products were visualized using 1.5% agarose gel electrophoresis under a UV transilluminator (Peiqing Science & Technology, Shanghai, China). Representative Sanger sequencing chromatograms are shown in Fig. 1: (A) homozygous wild-type genotype (CC), (B) heterozygous genotype (CT), and (C) homozygous mutant genotype (TT).

**Figure 1.**
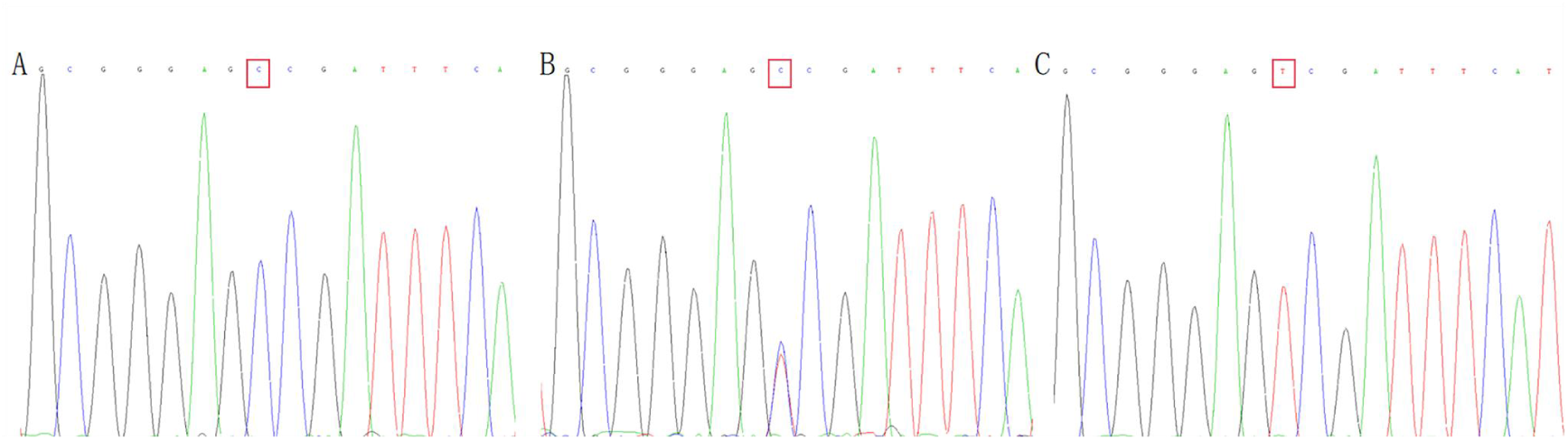
The Sanger sequencing chromatogram of *MTHFR*. (A)、(B) and (C) shows Sanger sequencing chromatogram of the homozygous wildtype genotype (CC) 、 heterozygous genotype(CT) and homozygous wildtype genotype (TT) respectly.

### 2.3 Methylation specific PCR (MSP)

Leukocyte DNA was subjected to bisulfite conversion using the TIANamp DNA Methylation Kit (Tiangen, Beijing, China) in accordance with the manufacturer’s protocol. Methylation-specific polymerase chain reaction (MSP) was performed using 100 ng of bisulfite-modified DNA with primers specific for the methylated or unmethylated sequences of the *MTHFR* promoter region. The primer sequences for the methylated allele were: forward 5′-AAGGAGGGGATATATAGTAAGAGGC-3′ and reverse 5′-CCTATTTTTAACAAAAATAAAACGTA-3′. The primer sequences for the unmethylated allele were:forward 5′-AGGAGGGGATATATAGTAAGAGGTG -3′ and reverse 5′-CCTATTTTTAACAAAAATAAAACATA-3′. PCR amplification was carried out under the following conditions: initial denaturation at 95 °C for 5 min; 35 cycles of denaturation at 94 °C for 20 s, annealing at 60 °C for 30 s, and extension at 72 °C for 20 s; followed by a final extension at 72 °C for 5 min. The 150 bp amplification products were separated by 2.0% agarose gel electrophoresis and visualized using a UV gel imaging system (Peiqing Science & Technology, Shanghai, China). As shown in Fig.2, a methylated band (M) indicated the presence of DNA methylation at the examined CpG sites, whereas an unmethylated band (U) indicated the absence of methylation at these loci.

**Figure 2:**
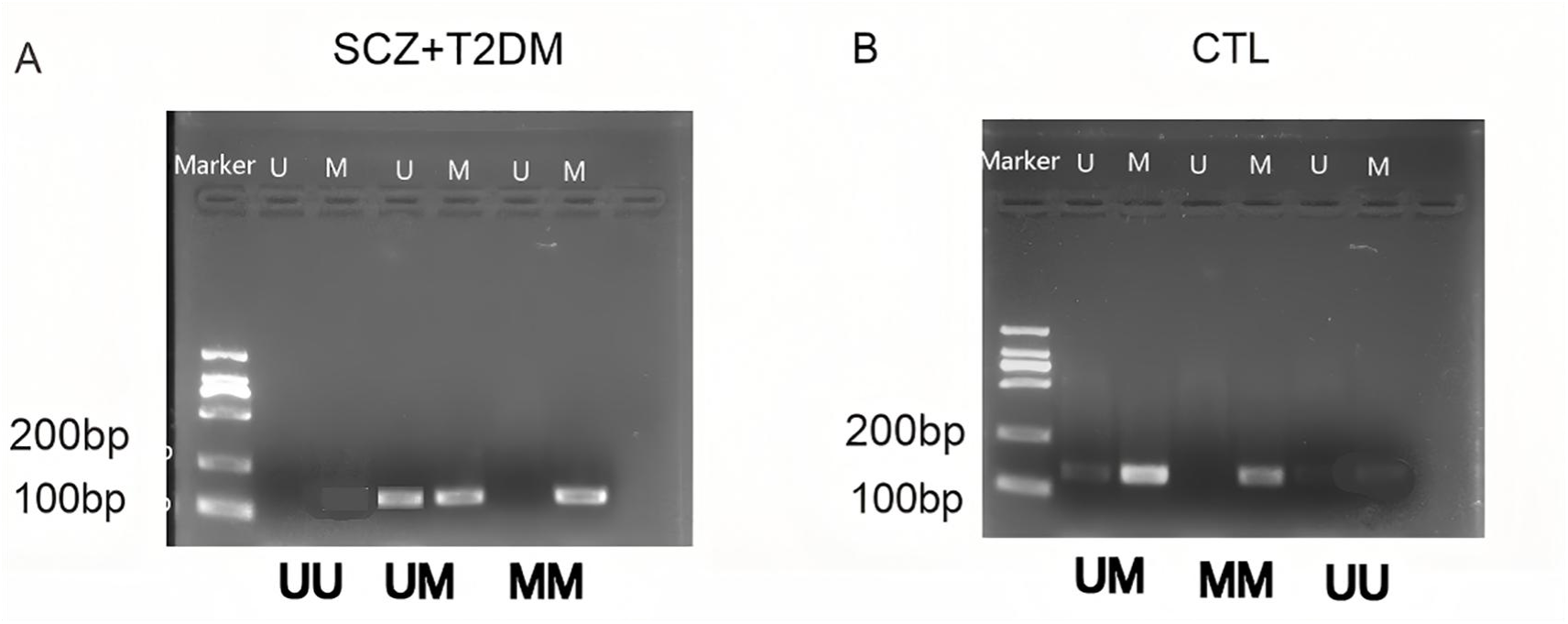
Methylation levels of the *MTHFR* promoter in peripheral blood from SCZ+T2DM patients and healthy controls. (A) Representative MSP results from the SCZ+T2DM group. (B) Representative MSP results from the control group. The bands were presented as M detected by methylated primer and as U detected by unmethylated primer.

### 2.4 Statistical analysis

Statistical analyses were performed using SPSS version 20.0 software. Hardy–Weinberg equilibrium was assessed using the chi-square (χ²) goodness-of-fit test. Categorical variables, including genotype and allele frequencies, were compared between groups using the chi-square test if the total sample size was ≥ 40 and all expected frequencies were ≥ 5. If these criteria were not met, the corrected chi-square test (for n ≥ 40 and 1 ≤ expected frequency < 5) or Fisher’s exact test (for n < 40 or expected frequency < 1) was applied instead. Quantitative data were presented as the mean ± standard deviation (SD) based on normality testing. Normally distributed data were compared using one- way analysis of variance (ANOVA) or independent-samples t-test, whereas non- normally distributed data were analyzed using the Kruskal–Wallis H test or Mann–Whitney U test. A P- value < 0.05 was considered statistically significant.

## 3 Results

### 3.1 Clinical characteristics

A total of 120 Han Chinese subjects were recruited in this study and assigned to four groups: the schizophrenia-only group, the type 2 diabetes mellitus (T2DM)-only group, the schizophrenia comorbid with T2DM group, and the healthy control group. No significant intergroup differences were observed in age, sex, educational level, smoking status, or drinking history (all *P*>0.05), suggesting that the four groups were well comparable (Table 1). By contrast, significant differences were detected in the histories of schizophrenia and T2DM among the groups (all *P*<0.05, Table 1).

**Table 1.** Clinical characteristics of the study subjects.

| Parameter | SCZ<br>(n=30) | T2DM<br>(n=30) | SCZ+T2DM<br>(n=30) | CTL<br>(n=30) | Statistic<br>value | *P* |
| --- | --- | --- | --- | --- | --- | --- |
| Age(years,<br>x±s) | 43.00±1.74 | 43.23±1.98 | 42.87±1.81 | 42.83±2.36 | *F*=0.250 | 0.861 |
| Gender(male/f<br>emale,n) | 17/13 | 16/14 | 20/10 | 21/9 | $\chi^2=2.397$ | 0.494 |
| Educational<br>level (≥high<br>school, n, %) | 14 (46.67) | 15 (50.00) | 13 (43.33) | 16 (53.33) | $\chi^2=0.667$ | 0.881 |
| Smoking<br>history (yes,<br>n, %) | 10(33.33) | 11(36.67) | 11(36.67) | 9(30.00) | $\chi^2=0.408$ | 0.939 |
| Drinking<br>history (yes,<br>n, %) | 9(30.00) | 9(30.00) | 10(33.33) | 8(26.67) | $\chi^2=0.317$ | 0.957 |
| Family history<br>of SCZ (yes,<br>n, %) | 6(20.00) | 0(0.00) | 4(13.33) | 1(3.33) | — | 0.023 |
| Family history<br>of T2DM<br>(yes, n, %) | 3(10.00) | 10(33.33) | 9(30.00) | 3(10.00) | $\chi^2=8.640$ | 0.034 |

*F*: One-way ANOVA; —: Fisher exact test ; $\chi^2$ : $\chi^2$ test; SCZ: schizophrenia; T2DM: type 2 diabetes
mellitus; SCZ+T2DM: schizophrenia patients with type 2 diabetes mellitus; CTL: healthy control.

### 3.2 *MTHFR* genotype and allelic distribution among the four groups

The chi-square (χ²) goodness-of-fit test revealed that the genotype distributions of the *MTHFR* rs1801133 polymorphism in all three patient groups and the control group were consistent with Hardy–Weinberg equilibrium (all *P*>0.05; Table 2).A significant association was identified between the *MTHFR* C677T (rs1801133) polymorphism and schizophrenia comorbid with type 2 diabetes mellitus (T2DM) in the overall study population. Both genotype and allele frequencies of the *MTHFR* rs1801133 variant differed significantly among the four groups: schizophrenia alone (SCZ), T2DM alone (T2DM), schizophrenia comorbid with T2DM (SCZ+T2DM), and healthy controls (CTL) (*P*=0.049; *χ²*=11.644, *P*=0.009; Table 3). These findings suggest that the *MTHFR* polymorphism may be implicated in the pathogenesis of schizophrenia and type 2 diabetes mellitus in the northern Han Chinese population.

**Table 2.** Analysis of *MTHFR* rs1801133 polymorphism for Hardy-Weinberg equilibrium.

| Group | Observed | | | Expected | | | n | $\chi^2$ | $P$ |
| --- | --- | --- | --- | --- | --- | --- | --- | --- | --- |
|  | CC | CT | TT | CC | CT | TT |  |  |  |
| SCZ | 7 | 11 | 12 | 5.21 | 14.58 | 10.21 | 30 | 1.8078 | 0.1788 |
| T2DM | 2 | 10 | 18 | 1.63 | 10.73 | 17.63 | 30 | 0.1414 | 0.7068 |
| SCZ+T2DM | 0 | 11 | 19 | 1.01 | 8.98 | 20.01 | 30 | — | 0.2183 |
| CTL | 6 | 12 | 12 | 4.80 | 14.40 | 10.80 | 30 | 0.8333 | 0.3611 |

—: Fisher exact test ; $\chi^2$ : $\chi^2$ test; *HWE*: Hardy–Weinberg equilibrium.

**Table 3.** Comparison of the *MTHFR* rs1801133 genotype and allele frequencies in four groups.

| Group | Genotypic Frequency | | | $\chi^2$ | $P$ | Allelic Frequency | | $\chi^2$ | $P$ |
| --- | --- | --- | --- | --- | --- | --- | --- | --- | --- |
|  | CC(%) | CT(%) | TT(%) |  |  | C(%) | T(%) |  |  |
| SCZ | 7(23.33) | 11(36.67) | 12(40.00) | — | 0.049 | 25(41.67) | 35(58.33) | 11.644 | 0.009 |
| T2DM | 2(6.67) | 10(33.33) | 18(60.00) |  |  | 14(23.33) | 46(76.67) |  |  |
| SCZ+T2DM | 0(0) | 11(36.67) | 19(63.33) |  |  | 11(18.33) | 49(81.67) |  |  |
| CTL | 6(20.00) | 12(40.00) | 12(40.00) |  |  | 24(40.00) | 36(60.00) |  |  |

—: Fisher exact test ; $\chi^2$ : $\chi^2$ test.

As shown in Table 4, the genotype distributions of the *MTHFR* C677T polymorphism differed significantly between patients with schizophrenia comorbid with type 2 diabetes mellitus (SCZ+T2DM) and healthy controls. In the SCZ+T2DM group, the frequencies of CC, CT, and TT genotypes were 0.00%, 36.67%, and 63.33%, respectively, compared with 20.00%, 40.00%, and 40.00% in the control group, with a statistically significant difference ( *P*=0.022). Accordingly, allele frequencies also differed significantly: the frequencies of the 677C and 677T alleles were 18.33% and 81.67% in the SCZ+T2DM group, versus 40.00% and 60.00% in the control group (*χ²*=6.817, *P*=0.009). These results demonstrate that the *MTHFR* C677T polymorphism is significantly associated with the risk of schizophrenia comorbid with type 2 diabetes mellitus. Notably, the frequency of the TT homozygous genotype was markedly higher in SCZ+T2DM patients than in healthy controls (63.33% vs. 40.00%), suggesting that the TT genotype of *MTHFR* rs1801133 may significantly increase susceptibility to this comorbid condition.

**Table 4.**
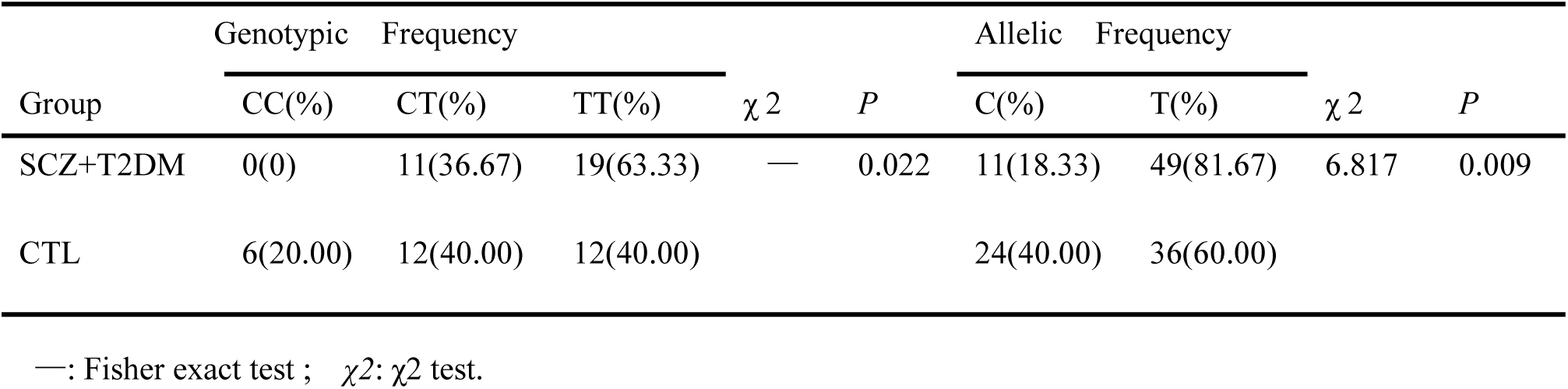
Comparison of the *MTHFR* rs1801133 genotype and allele frequencies between SCZ+T2DM and healthy controls.

### 3.3 Methylation levels of the *MTHFR* promoter in peripheral blood from SCZ+T2DM patients and healthy controls

To investigate the methylation status of the *MTHFR* promoter, peripheral blood samples from the SCZ+T2DM group and the control group were analyzed using methylation-specific PCR (MS- PCR; Fig. 2).The methylation status was classified as follows: full methylation (MM), indicated by a positive band only for methylated primers; full unmethylation (UU), indicated by a positive band only for unmethylated primers; and hemimethylation (UM), indicated by positive bands for both methylated and unmethylated primers. The overall methylation rate was calculated as (hemimethylation + full methylation) / total samples × 100%. As summarized in Table 5, the frequency of hemimethylation (UM) in the MTHFR promoter was 5.00% (1/20) in the SCZ+T2DM group and 30.00% (6/20) in the control group, with no statistically significant difference (*P*=1.000, OR=3.000, 95% CI: 0.122–73.642). In contrast, the frequency of full methylation (MM) was higher in the SCZ+T2DM group (90.00%, 18/20) than in the control group (60.00%, 12/20), although this difference was not statistically significant (*P*=0.781, O =0.333, 95% CI: 0.027–4.098). The combined methylation rate (UM + MM) was 95.00% (19/20) in the SCZ+T2DM group and 90.00% (18/20) in the control group, with no significant elevation in the comorbid group (*P*=1.000, OR=0.474, 95% CI: 0.039–5.688).

**Table 5.**
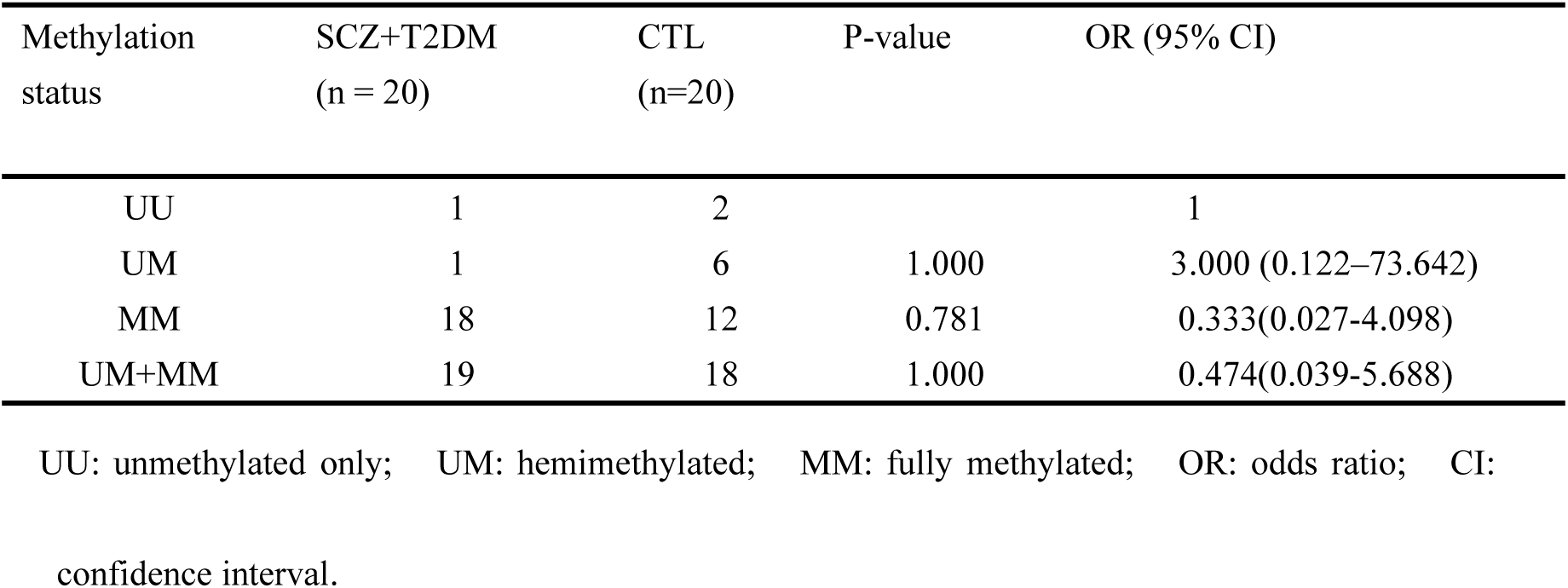
The frequency of *MTHFR* promoter methylation in peripheral blood from the SCZ+T2DM and control groups.

| Methylation status | SCZ+T2DM (n = 20) | CTL (n=20) | P-value | OR (95% CI) |
| --- | --- | --- | --- | --- |
| UU | 1 | 2 |  | 1 |
| UM | 1 | 6 | 1.000 | 3.000 (0.122–73.642) |
| MM | 18 | 12 | 0.781 | 0.333(0.027-4.098) |
| UM+MM | 19 | 18 | 1.000 | 0.474(0.039-5.688) |

UU: unmethylated only; UM: hemimethylated; MM: fully methylated; OR: odds ratio; CI: confidence interval.

## 4 Discussion

The comorbidity of schizophrenia (SCZ) and type 2 diabetes mellitus (T2DM) poses a complex clinical challenge, characterized by bidirectional pathogenic interactions, poor treatment outcomes, and increased healthcare burden[27]. Methylenetetrahydrofolate reductase (MTHFR), a key enzyme regulating folate metabolism and homocysteine (Hcy) homeostasis, has been independently implicated in the pathogenesis of both neuropsychiatric and metabolic disorders[28]. Prior to the present study, the differential roles of the *MTHFR* C677T polymorphism (rs1801133) and its promoter methylation in the development of SCZ-T2DM comorbidity remained elusive, particularly in the Han Chinese population. Thus, this study aimed to systematically explore the associations of *MTHFR* C677T polymorphism and promoter methylation with SCZ comorbid with T2DM in a Han Chinese cohort, thereby providing novel insights into the genetic and epigenetic mechanisms underlying this complex comorbid condition. The key results of this study demonstrated that the *MTHFR* C677T polymorphism was significantly associated with the risk of SCZ-T2DM comorbidity, whereas *MTHFR* promoter methylation showed no significant correlation with this comorbidity. Collectively, these findings indicate that genetic variation of *MTHFR* (C677T) may play a more predominant role than epigenetic modification (promoter methylation) in the pathogenesis of SCZ-T2DM comorbidity, especially in the Han Chinese population with unique genetic characteristics.

The present study provides robust and novel evidence that the *MTHFR* C677T polymorphism (rs1801133) is significantly associated with increased susceptibility to schizophrenia and type 2 diabetes (SCZ-T2DM) comorbidity in the Han Chinese population, aligning with our hypothesis that this functional genetic variant contributes to the co-occurrence of mental and metabolic disorders. Specifically, the frequencies of the T allele and TT genotype in the SCZ-T2DM comorbid group were significantly higher than those in the healthy control group, suggesting that the T allele may serve as a potential risk factor for the development of type 2 diabetes mellitus in patients with schizophrenia. This finding is biologically plausible and strongly supported by the well- characterized function of the *MTHFR* C677T polymorphism: the cytosine- to-thymine substitution at nucleotide position 677 leads to an alanine- to- valine missense mutation in the MTHFR protein, which markedly impairs its enzymatic stability and catalytic activity[17–18]. Compared with the wild-type CC genotype, the homozygous TT genotype has been shown to confer approximately a 75% decrease in MTHFR activity, whereas the heterozygous CT genotype results in an approximate 34% reduction[29]. Such compromised MTHFR function disrupts the one- carbon metabolism pathway, including folate cycle homeostasis and remethylation of homocysteine, thereby leading to sustained hyperhomocysteinemia and abnormal folate metabolism[28]. Hyperhomocysteinemia and folate insufficiency may exert synergistic detrimental effects on neuronal function, insulin signaling, glucose homeostasis, and systemic metabolic regulation, ultimately promoting the initiation and progression of SCZ-T2DM comorbidity[30]. Collectively, these results highlight the critical role of the *MTHFR* C677T polymorphism as a shared genetic risk factor underlying the comorbidity of schizophrenia and type 2 diabetes mellitus, and provide novel population-specific insights into the genetic mechanisms linking neuropsychiatric and metabolic diseases.

The potential mechanism underlying the association between the *MTHFR* C677T polymorphism and SCZ- T2DM comorbidity may be mediated through dysregulated one- carbon metabolism and subsequent metabolic and neurochemical abnormalities. On the one hand, elevated homocysteine (Hcy) levels resulting from reduced MTHFR activity have been consistently implicated in the pathogenesis of SCZ: increased Hcy can induce oxidative stress in neuronal cells, inhibit the synthesis of key neurotransmitters (such as dopamine and serotonin), and disrupt genomic DNA methylation patterns and DNA repair capacity, all of which represent core pathological features of SCZ[31]. A meta-analysis[32]conducted in Japanese populations has further validated a significant association between the *MTHFR* C677T polymorphism and SCZ susceptibility, while mendelian randomization analyses have suggested that increased plasma Hcy levels may increase the risk of SCZ. Nevertheless, it should be acknowledged that several studies have failed to replicate this association between the *MTHFR* C677T polymorphism and SCZ, which may be attributed to heterogeneity in study populations, sample sizes, and unadjusted confounding factors (such as psychotropic medication use and dietary folate intake). In the present study, we adjusted for multiple potential confounders including age, sex, educational level, smoking status, and drinking history, which considerably enhances the reliability and robustness of our findings. On the other hand, Hcy accumulation and impaired folate metabolism are also associated with the development of T2DM: elevated Hcy can induce insulin resistance by impairing pancreatic β- cell function and disrupting insulin signaling pathways[33], whereas folate insufficiency resulting from compromised MTHFR activity confers a higher susceptibility to T2DM. Consistent with these findings, a recent case-control study[34]have demonstrated that the *MTHFR* C677T polymorphism is associated with susceptibility to type 2 diabetes mellitus (T2DM) in Dali area population from Yunnan Province, China, with the TT genotype conferring a significantly elevated disease risk. Furthermore, the comorbidity of SCZ and T2DM may involve shared genetic and biological pathways, in which *MTHFR* serves as one of the key candidate genes. Collectively, these observations support the notion that the *MTHFR* C677T polymorphism contributes to the pathogenesis and progression of SCZ- T2DM comorbidity.

In contrast to the significant association of *MTHFR* C677T polymorphism with SCZ-T2DM comorbidity, our study found no significant difference in *MTHFR* promoter methylation levels between the SCZ-T2DM comorbid group and healthy control group. This result suggests that *MTHFR* promoter methylation may not be implicated in the pathogenesis of SCZ-T2DM comorbidity in the Han Chinese population, which is inconsistent with several previous studies focusing on single diseases. For example, a study in the Moroccan population[35]found that *MTHFR* promoter hypermethylation is associated with T2DM susceptibility, and other studies have suggested that *MTHFR* methylation may be involved in the development of SCZ[36]. The discrepancy between our findings and previous studies may be explained by the following factors. First, previous studies focused on single diseases (SCZ or T2DM) or their complications, while our study focused on the comorbidity of the two diseases. It is possible that *MTHFR* promoter methylation is involved in the progression of single diseases or their complications, but not in the occurrence of comorbidity. Second, the targeted region of *MTHFR* promoter methylation may vary across studies. We detected the core CpG island of the *MTHFR* promoter, while other studies may have targeted different regions, leading to divergent results. Third, the sample size and population characteristics may also influence the outcomes. Our study was conducted in a Han Chinese cohort, while previous studies may have included populations of different ethnicities, and *MTHFR* methylation profiles may vary across ethnic groups. Additionally, the regulatory effect of *MTHFR* promoter methylation on glucose metabolism may be offset by other factors, such as antipsychotic medication use and lifestyle factors.

This study has several limitations that should be addressed in future research. First, the sample size is limited to a Han Chinese cohort, which restricts the generalizability of the results to other ethnic groups, given the population-specific differences in *MTHFR* genetic and epigenetic patterns. Second, we only interrogated the *MTHFR* C677T polymorphism and the core CpG island of the promoter, and did not explore other polymorphisms (e.g: A1298C, rs1801131) or methylation sites of *MTHFR*, which may miss other potential associations. Third, we did not measure MTHFR enzyme activity and folate levels, which may limit the exploration of the underlying mechanism. Fourth, MSP has inherent limitations: as a qualitative rather than quantitative assay, it fails to accurately quantify methylation levels and capture subtle variations. Additionally, the cross-sectional design of the study restricts the ability to draw conclusions about causality. Future studies should expand to multi-ethnic cohorts, integrate multi-omics data (genetics, epigenetics, transcriptomics), and conduct longitudinal follow-ups to validate the findings. We further recommend that bisulfite sequencing PCR (BSP) be employed instead of MSP for more accurate and comprehensive methylation analysis. Large-sample, multi- center, and prospective study designs are also warranted to further clarify the epigenetic role of *MTHFR* in the pathogenesis of this comorbid condition.

## 5 Conclusion

In conclusion, our study demonstrates that the *MTHFR* C677T polymorphism is significantly associated with SCZ-T2DM comorbidity in the Han Chinese population, while *MTHFR* promoter methylation has no significant correlation with this comorbidity. These results suggest that genetic variation in *MTHFR* plays a more important role than promoter methylation in the comorbidity of SCZ and T2DM. Our findings provide new insights into the genetic basis of SCZ-T2DM comorbidity and may contribute to the development of personalized prevention and treatment strategies for this comorbidity in the Han Chinese population.

## Data Availability

The data that support the findings of this study are available from the corresponding author upon reasonable request.

## 6 List of abbreviations

SCZ: schizophrenia
T2DM: type 2 diabetes mellitus
MTHFR: methylenetetrahydrofolate reductase
MSP: methylation-specific PCR
HWE: Hardy-Weinberg equilibrium
DR: diabetic retinopathy
DN: diabetic nephropathy
DPN: diabetic peripheral neuropathy
SNP: single-nucleotide polymorphism
IDF: International Diabetes Federation
DSM-5: Diagnostic and Statistical Manual of Mental Disorders, 5th edition
FPG: fasting plasma glucose
2hPG: 2-hour Postprandial Glucose
BSP: bisulfite sequencing PCR

## 7 Conflict of Interest

The authors declare that the research was conducted in the absence of any commercial or financial relationships that could be construed as a potential conflict of interest.

## 8 Author Contributions

CCY: Investigation, Data curation, Writing – original draft. ZJ: Funding acquisition, Methodology, Validation. RZ: Investigation, Methodology, Formal analysis. RRL: Investigation, Methodology, Formal analysis. XDJ: Project administration, Resources, Writing – review & editing. FJY: Supervision, Writing – review & editing, Conceptualization. XZW: Data curation, Validation, Supervision. KL: Data curation, Validation, Supervision.

## 9 Funding

This project was supported by the Affiliated Hospital (Teaching Hospital) Research and Development Foundation of Shandong Second Medical University (2024FYM133, 2025FYM133 and 2025FYM139). Policy-Guided Scientific and Technological Projects of Key R&D Plan in Liaocheng City (2024YD41).

## 10 Acknowledgements

The authors thank for all patients and healthy subjects for their participation in the study, and FJY, XDJ and clinical researchers from Liaocheng People’s Hospital of Joint Laboratory for Translational Medicine Research for their supports and help.

## Notes

### Competing Interest Statement

The authors have declared no competing interest.

### Author Declarations

The study protocol was approved by the Institutional Ethics Committee of Liaocheng Fourth People's Hospital.

## References

1. Jauhar S, Johnstone M, McKenna PJ. Schizophrenia. Lancet. 2022;399(10323):473–86. doi: 10.1016/s0140-6736(21)01730-x.

2. GBD 2019 Mental Disorders Collaborators. Global, regional, and national burden of 12 mental disorders in 204 countries and territories, 1990-2019: a systematic analysis for the Global Burden of Disease Study 2019. Lancet Psychiatry. 2022;9(2):137–50. doi: 10.1016/s2215-0366(21)00395-3.

3. Velligan DI, Rao S. The Epidemiology and Global Burden of Schizophrenia. J Clin Psychiatry. 2023;84(1):MS21078COM5.doi: 10.4088/jcp.Ms21078com5.

4. Huang Y, Wang Y, Wang H, Liu Z, Yu X, Yan J, et al. Prevalence of mental disorders in China: a cross-sectional epidemiological study. Lancet Psychiatry. 2019;6(3):211–24. doi: 10.1016/s2215-0366(18)30511-x.

5. Solmi M, Seitidis G, Mavridis D, Correll CU, Dragioti E, Guimond S, et al. Incidence, prevalence, and global burden of schizophrenia - data, with critical appraisal, from the Global Burden of Disease (GBD) 2019. Mol Psychiatry. 2023;28(12):5319–27. doi: 10.1038/s41380-023-02138-4.

6. Kadakia A, Catillon M, Fan Q, Williams GR, Marden JR, Anderson A, et al. The Economic Burden of Schizophrenia in the United States. J Clin Psychiatry. 2022;83(6):22m14458. doi: 10.4088/JCP.22m14458.

7. Martins R, Kadakia A, Williams GR, Milanovic S, Connolly MP. The Lifetime Burden of Schizophrenia as Estimated by a Government-Centric Fiscal Analytic Framework. J Clin Psychiatry. 2023;84(5):22m14746. doi: 10.4088/JCP.22m14746.

8. Majumdar S, Sinha B, Dastidar BG, Gangopadhyay KK, Ghoshal S, Mukherjee JJ, et al. Assessing prevalence and predictors of depression in Type 2 Diabetes Mellitus (DM) patients - The DEPDIAB study. Diabetes Res Clin Pract. 2021;178:108980. doi: 10.1016/j.diabres.2021.108980.

9. Luo A, Xie Z, Wang Y, Wang X, Li S, Yan J, et al. Type 2 diabetes mellitus-associated cognitive dysfunction: Advances in potential mechanisms and therapies. Neurosci Biobehav Rev. 2022;137:104642. doi: 10.1016/j.neubiorev.2022.104642.

10. Majety P, Lozada Orquera FA, Edem D, Hamdy O. Pharmacological approaches to the prevention of type 2 diabetes mellitus. Front Endocrinol (Lausanne). 2023;14:1118848. doi: 10.3389/fendo.2023.1118848.

11. Singh A, Shadangi S, Gupta PK, , Rana S. Type 2 Diabetes Mellitus: A Comprehensive Review of Pathophysiology, Comorbidities, and Emerging Therapies. Compr Physiol. 2025;15(1):e70003. doi: 10.1002/cph4.70003.

12. Saeedi P, Petersohn I, Salpea P, Malanda B, Karuranga S, Unwin N, et al. Global and regional diabetes prevalence estimates for 2019 and projections for 2030 and 2045: Results from the International Diabetes Federation Diabetes Atlas, 9(th) edition. Diabetes Res Clin Pract. 2019;157:107843. doi: 10.1016/j.diabres.2019.107843.

13. Li Y, Teng D, Shi X, Qin G, Qin Y, Quan H, et al. Prevalence of diabetes recorded in mainland China using 2018 diagnostic criteria from the American Diabetes Association: national cross sectional study. BMJ. 2020;369:m997. doi: 10.1136/bmj.m997.

14. Dong K, Wang S, Qu C, Zheng K, Sun P. Schizophrenia and type 2 diabetes risk: a systematic review and meta-analysis. Front Endocrinol (Lausanne). 2024;15:1395771. doi: 10.3389/fendo.2024.1395771.

15. Bagher AM, Alkhaldi MF, Somaily JA, Altheyab DA, Khafaji MA, Awad RF, et al. Methylenetetrahydrofolate reductase (MTHFR) C677T and A1298C polymorphisms are associated with major depressive disorder in the Saudi patients attending Erada complex for mental health and Erada services - Jeddah, Saudi Arabia. Pharmazie. 2024;79(10):228–32. doi:10.1691/ph.2024.4601

16. Sugijo H, Sargowo D, Widjajanto E, Romdoni R. The role of methylenetetrahydrofolate reductase C677T gene polymorphism as a risk factor for coronary artery disease: a cross-sectional study in the Sidoarjo Regional General Hospital. Pan Afr Med J. 2022;41:212. doi: 10.11604/pamj.2022.41.212.24916.

17. Mohammed NO, Ali IA, Elamin BK, Saeed BO. 5,10-methylenetetrahydrofolate reductase C677T gene polymorphism as a risk factor for premature coronary artery disease in patients with type 2 diabetes mellitus. Front Endocrinol (Lausanne). 2024;15:1502497. doi: 10.3389/fendo.2024.1502497.

18. Pathak D, Shrivastav D, Verma AK, Alsayegh AA, Yadav P, Khan NH, et al. Role of metabolizing MTHFR gene polymorphism (rs1801133) and its mRNA expression among Type 2 Diabetes. J Diabetes Metab Disord. 2022;21(1):511–6. doi: 10.1007/s40200-022-01001-7.

19. Zhang YX, Yang LP, Gai C, Cheng C-C, Guo Z-y, Sun H-M, et al. Association between variants of MTHFR genes and psychiatric disorders: A meta-analysis. Front Psychiatry. 2022;13:976428. doi: 10.3389/fpsyt.2022.976428.

20. Meng Y, Liu X, Ma K, Zhang L, Lu M, Zhao M, et al. Association of MTHFR C677T polymorphism and type 2 diabetes mellitus (T2DM) susceptibility. Mol Genet Genomic Med. 2019;7(12):e1020. doi: 10.1002/mgg3.1020.

21. Kaimala S, Yassin LK, Hamad MIK, Allouh MZ, Sampath P, AlKaabi J, et al. Epigenetic crossroads in metabolic and cardiovascular health: the role of DNA methylation in type 2 diabetes and cardiovascular diseases. Cardiovasc Diabetol. 2025;24(1):231. doi: 10.1186/s12933-025-02800-x.

22. Ghattas M, El-Shaarawy F, Mesbah N, Abo-Elmatty D. DNA methylation status of the methylenetetrahydrofolate reductase gene promoter in peripheral blood of end-stage renal disease patients. Mol Biol Rep. 2014;41(2):683–688. doi:10.1007/s11033-013-2906-7

23. Osunkalu VO, Taiwo IA, Makwe CC, Abiola AA, Quao RA, Anorlu RI. Epigenetic Modification in Methylene Tetrahydrofolate Reductase (MTHFR) Gene of Women with Pre-eclampsia. J Obstet Gynaecol India. 2021;71(1):52–57. doi: 10.1007/s13224-020-01374-w.

24. Cai C, Meng C, He S, Gu C, Lhamo T, Draga D, et al. DNA methylation in diabetic retinopathy: pathogenetic role and potential therapeutic targets. Cell Biosci. 2022;12(1):186. doi: 10.1186/s13578-022-00927-y.

25. Deng G, Feng Z, Kong X, Gao P, Jiang Y, Liu Y, et al. Association Between DNA Methylation of MTHFR and Diabetic Kidney Disease. J Diabetes Res. 2025;2025:8096423. doi: 10.1155/jdr/8096423.

26. First MB. Diagnostic and statistical manual of mental disorders, 5th edition, and clinical utility. J Nerv Ment Dis. 2013;201(9):727–9. doi: 10.1097/NMD.0b013e3182a2168a.

27. Mizuki Y, Sakamoto S, Okahisa Y, Yada Y, Hashimoto N, Takaki M, et al. Mechanisms Underlying the Comorbidity of Schizophrenia and Type 2 Diabetes Mellitus. Int J Neuropsychopharmacol. 2021;24(5):367–382. doi: 10.1093/ijnp/pyaa097.

28. Araszkiewicz AF, Jańczak K, Wójcik P, Białecki B, Kubiak S, Szczechowski M, et al. MTHFR Gene Polymorphisms: A Single Gene with Wide-Ranging Clinical Implications-A Review. Genes (Basel). 2025;16(4):441. doi: 10.3390/genes16040441.

29. Wan L, Li Y, Zhang Z, Sun Z, He Y, Li R. Methylenetetrahydrofolate reductase and psychiatric diseases. Transl Psychiatry. 2018;8(1):242. doi: 10.1038/s41398-018-0276-6.

30. Borovcanin MM, Vesic K, Petrovic I, Jovanovic IP, Mijailović NR. Diabetes mellitus type 2 as an underlying, comorbid or consequent state of mental disorders. World J Diabetes. 2023;14(5):481–493.doi: 10.4239/wjd.v14.i5.481.

31. Yang X, Yang H, Li N, Li C, Liang W, Zhang X. Increased serum homocysteine in first episode and drug-naïve individuals with schizophrenia: sex differences and correlations with clinical symptoms. BMC Psychiatry. 2022;22(1):759. doi: 10.1186/s12888-022-04416-x.

32. Nishi A, Numata S, Tajima A, Kinoshita M, Kikuchi K, Shimodera S, et al. Meta-analyses of blood homocysteine levels for gender and genetic association studies of the MTHFR C677T polymorphism in schizophrenia. Schizophr Bull. 2014;40(5):1154–1163. doi: 10.1093/schbul/sbt154.

33. Zhang X, Qu YY, Liu L, Qiao YN, Geng HR, Lin Y, et al. Homocysteine inhibits pro-insulin receptor cleavage and causes insulin resistance via protein cysteine-homocysteinylation. Cell Rep. 2021;37(2):109821. doi: 10.1016/j.celrep.2021.109821.

34. Liu Y, Pu G, Yang C, Wang Y, Jin K, Wang S, et al. Association analysis of MTHFR (rs1801133 and rs1801131) gene polymorphism towards the development of type 2 diabetes mellitus in Dali area population from Yunnan Province, China. PeerJ .2024;12:e18334. doi: 10.7717/peerj.18334.

35. El Alami H, Bouqdayr M, Errafii K, Messaoudi N, Sehli S, Al Idrissi N, et al. Association of CpG site of MTHFR gene promoter and type 2 diabetes in Moroccan population susceptibility. Nucleosides Nucleotides Nucleic Acids. Published online July 10, 2025. doi:10.1080/15257770.2025.2532089.

36. Wan L, Zhang G, Liu M, Wang C, Li Y, Li R. Sex-specific effects of methylenetetrahydrofolate reductase polymorphisms on schizophrenia with methylation changes. Compr Psychiatry. 2019;94:152121. doi: 10.1016/j.comppsych.2019.152121.

